# Impact of γ-herpes viruses’ status on established risk factor profile of oral squamous cell carcinoma tissues in a group of Sri Lankan male

**DOI:** 10.1101/2023.08.02.23293533

**Authors:** Manosha Lakmali Perera, Irosha Rukmali Perera

## Abstract

Oral cancer with multifactorial aetiology was established as a public health challenge globally especially locally. Alarmingly, oral cancer ranks 1st among Sri Lankan males and 8^th^ among Sri Lankan females with 2-3 death daily. Oral squamous cell carcinoma (OSCC) accounts for 90-95% of oral malignancies in most countries. Perhaps, infectious agents are responsible for approximately 20% of global cancer incidence. Among them, Epstein-Barr virus (EBV) and Kaposi’s sarcoma-associated herpes virus (KHSV) are γ-herpes viruses linked with mucosal malignancies also. There is a dearth of information on oncogenic viral infections in oral cancer patients. Thus, the present study aimed to investigate the impact of γ-herpes viruses’ status on the -established risk factor profile of OSCC tissue in a group of Sri Lankan male patients. Ethical approvals were obtained from Sri Lanka and Australia. A large unmatched case-control study was conducted on Oral and Maxillo-Facial Surgical (OMF) units located in six provinces. Consequently, a representative sub-sample of histopathologically confirmed 29 OSCC cases and clinically diagnosed 25 Fibroepithelial-poly (FEP)controls was selected from the main, adhering to stringent inclusion criteria, for cases at least with betel quid chewing, while excluding none of oral risk habits. Incisional biopsies of cases and excisional biopsies of controls were collected, transported, stored, and dispatched as frozen tissues at - 80^0^ C. Then, DNA was extracted from frozen specimens. Subsequently, real-time PCR was performed to detect γ-herpes viruses separately. Socio demographic and clinical data were obtained entered and analysed using SPSS-21 Statistical Package. Descriptive statistics and Fisher’s exact test were used to compare groups ((cell counts <5). The overall EBV prevalence was calculated as 34 (64.2%). In OSCC cases the EBV positivity was higher 21(77.8%) than the FEP controls 13 (50.0%), and this difference was statistically significant (p<0.05). Neither cases nor controls infected with KSHV/HHV-8. In oral risk habit comparison, the cases were better substance abusers, especially in terms of betel quid chewing and alcohol consumption than the control group (p<0.05), The impacts of γ-herpes viruses’ status on established risk factors are noteworthy to investigate by adequately powered prospective studies exclusively in local and regional context.

## Introduction

Nearly, 13% of global malignancies are considered as with viral aetiology depending on geographic and population specificity as reservoirs and mode of transmission of these oncogenic viruses may be due to life style related risk factors [1]. Chronic persistence of viral genome in the host is a predisposing factor for viral cause of cancers [2]. Among the oncogenic viruses two members of Herpesviridae family as well as Gammaherpesvirinae subfamily Epstein-Barr virus (EBV; and human herpesvirus 8 (HHV-8) gained much attention as their role in aetiology of oral cancer yet to be revealed. The EBV also known as HHV-4 is the main cause of nasopharyngeal cancers, a distinct biological entity [3,4]. The other member of the same subfamily KSHV /HHV-8 is the aetiological agent of Kaposi’s sarcoma in oral and extra-oral sites prevalent in immunosuppressed individuals, especially in patients with acquired immune deficiency syndrome (AIDS). There is substantial evidence of the association of EBV with oral cancer. The impact of EBV and KSHV on the established risk factor profile is yet to be discovered. The present study intended to investigate the impact of □viruses’ status on the established risk profile of oral squamous cell carcinoma tissues in a group of Sri Lankan male patients.

## Methodology

A sub sample from a powered retrospective study was selected to represent the vast majority of OSCC patients as described previously [4]. Each participant provided a written consent. Ethical approval for this study was obtained by the University of Peradeniya, Sri Lanka (FRC/ FDS/UOP/E/2014/32), and the Griffith University Human Research Ethics Committee, Australia (DOH/18/14/HREC).Deep tissue samples (∼100 mg each) were dissected from fresh incisional biopsies for OSCC cases and excisional biopsies for FEP stored at -80^0^C, avoiding contamination from the tumor surface. DNA was extracted with the Gentra Puregene Tissue kit (Qiagen) strictly adhering to manufacturer’s protocol for solid tissue. [Cat no. 158689] as described previously [3,4]. The quality of extracted DNA was assessed by β-globin PCR with the primers PCO3 and PCO4 [4].The rt PCR assay was set up to amplify 106 bp of EBV from primer sequences as described previously [3, 4]. A 10-μl volume of each test sample was placed into the wells of a white 384 -well plate in an EppendorfepMotion 5075. rt PCR was then performed using a Quant 6-real -time machine as described previously. Absence of PCR inhibitors in extracted DNA was assessed by β-globin PCR with the primers PCO3 and PCO4 [4].The real time PCR assay was set up to amplify 106 bp of HHV-8, from previously published primer sequences as described earlier [4]. The data were entered and analysed using SPSS-21 Statistical Package. Descriptive statistics and Fisher’s exact test were used to compare groups ((cell counts <5).

## Results and Discussion

**Table 01:** Distribution of the Cases and Controls by Established Risk Habit Profile.

| Variable | Cases n=29 | Controls n=25 | p-value |  |
| --- | --- | --- | --- | --- |
|  | N % | N % |  |  |
| *Betel Chewing Habit* |  |  |  |  |
| Never | 0 (0.0) | 4(16.0) | 0.003* | (p<0.05) |
| Past | 5 (20.0) | 2(8.0) |  |  |
| Sometimes | 1(4.0) | 7 (28.0) |  |  |
| Daily | 23(76.0) | 12 (48.0) |  |  |
| **Total** | **29 (100.0)** | **25 (100.0)** |  |  |
| *Smoking Habit* |  |  |  |  |
| Never | 5 (17.2) | 8(32.0) | 0.418* | (p>0.05) |
| Past | 8 (27.6) | 6(24.0) |  |  |
| Sometimes | 4 (13.8) | 5(20.0) |  |  |
| Daily | 12(41.4) | 6 (24.0) |  |  |
| **Total** | **29 (100.0)** | **25 (100.0)** |  |  |
| *Alcohol Consumption* |  |  |  |  |
| Never | 3 (10.3) | 3(12.0) | 0.001* | (p<0.05) |
| Past | 6(20.7) | 2(8.0) |  |  |
| Sometimes | 4(13.8) | 16 (64.0) |  |  |
| Weekly | 16 (55.2) | 4 (16.0) |  |  |
| **Total** | **29 (100.0)** | **25 (100.0)** |  |  |

**Table 02:** Distribution of Cases and Controls by EBV Status.

| HBV Status | Cases n=29 |  | Controls n=25 |  | Total | p-value |
| --- | --- | --- | --- | --- | --- | --- |
|  | N | (%) | N | (%) | N (%) |  |
| Positive | 21 | (77.8) | 13 | (50.0) | 34(64.2) | 0.035*(p<0.05) |
| Negative | 06 | (22.2) | 13 | (50.0) | 19(35.8) |  |
| **Total** | **27** | **(100.0)** | **26** | **(100.0)** | **53(100.0)** |  |

\* Chi-square Test of Statistical Significance

**Table 03:** Distribution of Cases and Controls by KSHV status.

| KSHV Status | Cases n=29 |  | Controls n=25 |  | Total | p-value |
| --- | --- | --- | --- | --- | --- | --- |
|  | N | (%) | N | (%) | N (%) |  |
| Positive | 0 | (0.00) | 0 | (0.00) | 0 (0.00) | - |
| Negative | 27 | (100.0) | 26 | (100.0) | 0 (0.00) |  |
| **Total** | **27** | **(100.0)** | **26** | **(100.0)** | **53(100.0)** |  |

EBV prevalence in OSCC cases vary from 0-100% depending on the population and the location. EBV positivity of OSCC cases was much higher (77.8%) than (35.0%) in the past study conducted by Jalouli and colleagues [4]. Consistently, increased EBV prevalence in OSCC cases was found in Taiwan (82.5%), Japan (76.6%), Yamen (73.3%), and Hungary (73.8%) [4]. KSHV was not detected in either OSCC cases or FEP controls. In oral risk, habit comparison all cases (100%) but controls (84%) were present or betel quid chewers. The difference between the two groups was statistically significant (p<0.05). With regards to alcohol consumption, the majority (55.2%) of cases were regular drinkers whereas the majority (64.0%) of controls was occasional drinkers. Despite both groups being alcohol consumers, cases reported significantly higher weekly drinking of alcohol (p< 0.05). Corroborating the finding of the present study, nearly 2-fold higher odds ratio was found in the Shammah users/EBV-positive group compared to the Shammah users/EBV-negative group and Shammah is a type of smokeless tobacco product used in Yamen [4]. The small sample size is a limitation in this study.

## Conclusion

EBV might have an impact on betel quid chewing and alcohol consumption Thus, adequately powered prospective studies are recommended to find-out the significance of our conclusion.

## Data Availability

All data produced in the present study are available upon reasonable request to the authors

